# Multi-ancestry genome-wide association meta-analysis of hepatocellular carcinoma identifies eight novel risk genes including *MYC*, *MAP3K9*, *DHRS1*, and *MTTP*

**DOI:** 10.64898/2025.12.16.25342341

**Authors:** I Chinaka, A Schofield, CI Amos, R Lewis, VL Chen, Y Han, M Hassan, S Shetty, JP Mann

**Affiliations:** Department of Medicine, St James’ Hospital, Dublin, Ireland; Department of Immunology & Immunotherapy, School of Infection, Inflammation and Immunology, College of Medicine and Health, University of Birmingham, UK; Department of Internal Medicine, University of New Mexico, Albuquerque, NM, USA; Division of Gastroenterology and Hepatology, Mayo Clinic, Rochester, Minnesota, USA.; Division of Gastroenterology and Hepatology, Department of Internal Medicine, University of Michigan, Ann Arbor, MI, USA; Department of Epidemiology, The University of Texas MD Anderson Cancer Center, Houston, Texas, USA; Liver Unit (including small bowel transplantation), Birmingham Women’s and Children’s NHS Foundation Trust, Birmingham, UK

**Author notes:** **Correspondence**: Dr. Jake P. Mann Centre for Liver and gastroenterology research, Department of Immunology & Immunotherapy, School of Infection, Inflammation and Immunology, College of Medicine and Health, University of Birmingham, UK.

## Abstract

**Background & Aims:** Hepatocellular carcinoma (HCC) is the third top cause of cancer death globally, often arising on a background of cirrhosis. Here, we aimed to establish novel genetic drivers of HCC across ancestries at a population level through a large meta-analysis of all-cause HCC.

**Approach & Results:** We included 15 cohorts comprising 17,329 HCC cases and 2,424,298 controls in this meta-analysis. We found 15 genome-wide significant (P < 5x10^-8^) germline loci, 6 novel in/near *GCKR*, *MTTP*, *ADH5*, *MYC*, *MAP3K9*, and *GABPB2*. *MAP3K9*, *TERT*, and *GABPB2* variants act independently of cirrhosis on both co-localisation analysis and sensitivity analyses. There was significant ancestral heterogeneity in 6 loci including variants in the HLA locus that had divergent effects on HCC risk between East Asian and European ancestries. Fine-mapping identified 11 potentially causal coding variants, including p.Leu446Pro in GCKR and p.Asp418Glu in MEN1. *MEN1*, *MYC*, and *TERT* are all involved in the beta-catenin pathway transactivation complex. Transcriptome-wide analysis identified enrichment of germline-encoded *DHRS1* in HCC. Regulome-wide analysis replicated the germline signal for *EPHA2* and found a novel chromatin accessible region containing genes *ZNF367* and *HABP4*. Finally, we demonstrated that population-level genetic architecture for HCC overlaps with steatotic and viral liver disease, and individuals with genetic risk for lower BMI have higher risk of HCC.

**Conclusions:** Genetic risk for HCC is determined by germline susceptibility to beta-catenin pathway activation and cirrhosis. HCC is driven by both heterogenous and homogenous genetic factors across ancestries, which require further ancestral diversity in liver GWAS to fully dissect.

## Introduction

Hepatocellular carcinoma (HCC) is the 3^rd^ top cause of cancer death globally(1). It typically arises on a background of cirrhosis, which can be caused by any aetiology of liver disease(2). HCC may also develop in non-cirrhotic individuals, particularly in chronic hepatitis B virus (HBV) infection and in metabolic dysfunction-associated steatotic liver disease (MASLD). Identification of genetic risk factors is critical to mitigate rapidly growing burden of HCC(3).

Several previous genome-wide association studies (GWAS) have discovered a total of 35 independent risk loci in individuals of European and East Asian genetic ancestry(4–7).

Recent GWAS have demonstrated that combining aetiologies of liver disease can increase power to detect variants that perturb common pathways in the mechanisms of liver fibro-inflammation(8). For example, an all-cause cirrhosis GWAS identified p.Thr165Pro in MTARC1 as a protective variant, which was not apparent in single-aetiology analyses(9).

Two GWAS meta-analyses of HCC have expanded our understanding of germline variation that drives HCC(6,7). Genes that contribute to HCC can be broadly divided into those that act via cirrhosis and those that are independent of liver fibrosis. Variants in *TM6SF2* and *PNPLA3* are the two most replicated genes that increase risk of cirrhosis liver fibrosis, primarily via increasing hepatic lipid accumulation(10). Whereas, variants in *TERT* appear to directly affect HCC development without any mediation via chronic liver disease related pathways(7,11).

Here, we performed a multi-ancestry meta-analysis (MAMA) of HCC GWAS, which included transcriptome-wide and regulome-wide association analyses. In doing so, we identified eight novel loci as putative risk genes for HCC and established a baseline for future studies with inclusion of diverse ancestries.

## Results

### GWAS meta-analyses identify 9 known and 6 novel significant genes

We used 4 datasets from East Asian, 7 European, 1 African American, 1 Admixed American, and 2 mixed ancestry cohorts (Fig 1 and Supplementary Table 1). Overall (including replication cohort), this included 17,329 HCC cases and 2,424,298 controls in the MAMA. There was no evidence of overlap between cohorts within ancestries based on linkage disequilibrium (LD) score regression (LDSC), which were close to zero or within the 95% confidence interval (Supplementary Table 2).

**Fig 1.**
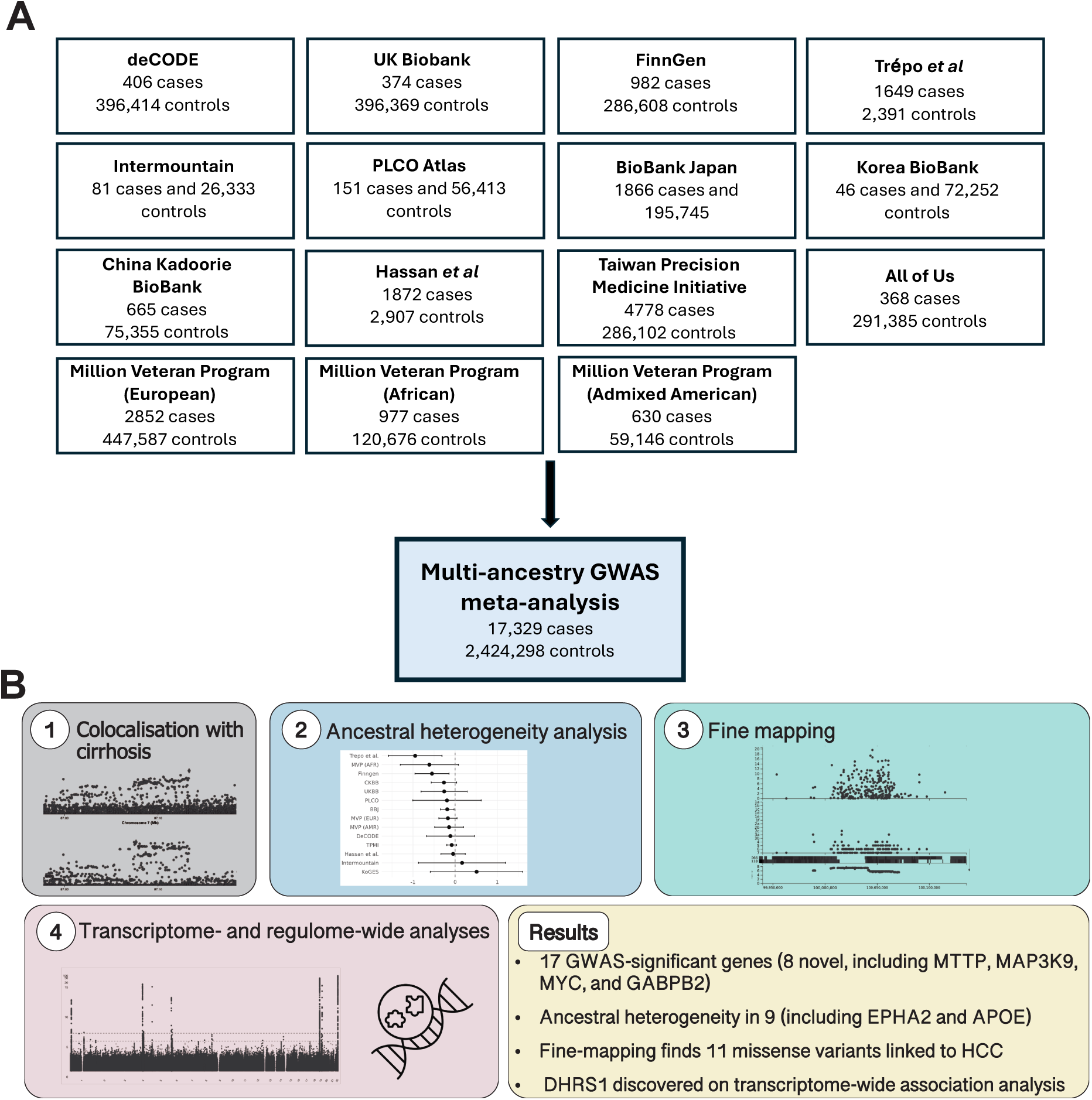
Study overview. (A) 15 cohorts from four genetic ancestries were included in the meta-analysis, totalling 17,329 hepatocellular carcinoma (HCC) cases. (B) Post-GWAS analysis included colocalisation with cirrhosis, fine mapping, and transcriptome-/regulome-wide association studies.

Following data harmonization and mapping to genome build hg38, MAMA was performed using meta-regression of multi-ethnic genetic association (MR-MEGA) and METAL (including single-ancestry analyses for European and East Asian cohorts). As previously described, MR-MEGA has greater power to detect heterogenous effects across different cohorts as well as distinguishing variation in effect estimates from ancestry-level genetic variation. Whereas METAL had better power for identification of homogenous allelic effects. MR-MEGA also facilitates calculation of posterior probabilities for fine mapping across ancestries using allele frequencies in the different cohorts.

In combination, we found 5 novel and 9 known loci that met GWAS significance of P < 5x10^-8^ (Table 1, Fig. 2 and Supplementary Figures 1 & 2, and Supplementary Tables 3 & 4). Out of 14 total GWAS-significant loci, 9 were observed across both MR-MEGA and METAL models.

**Fig 2.**
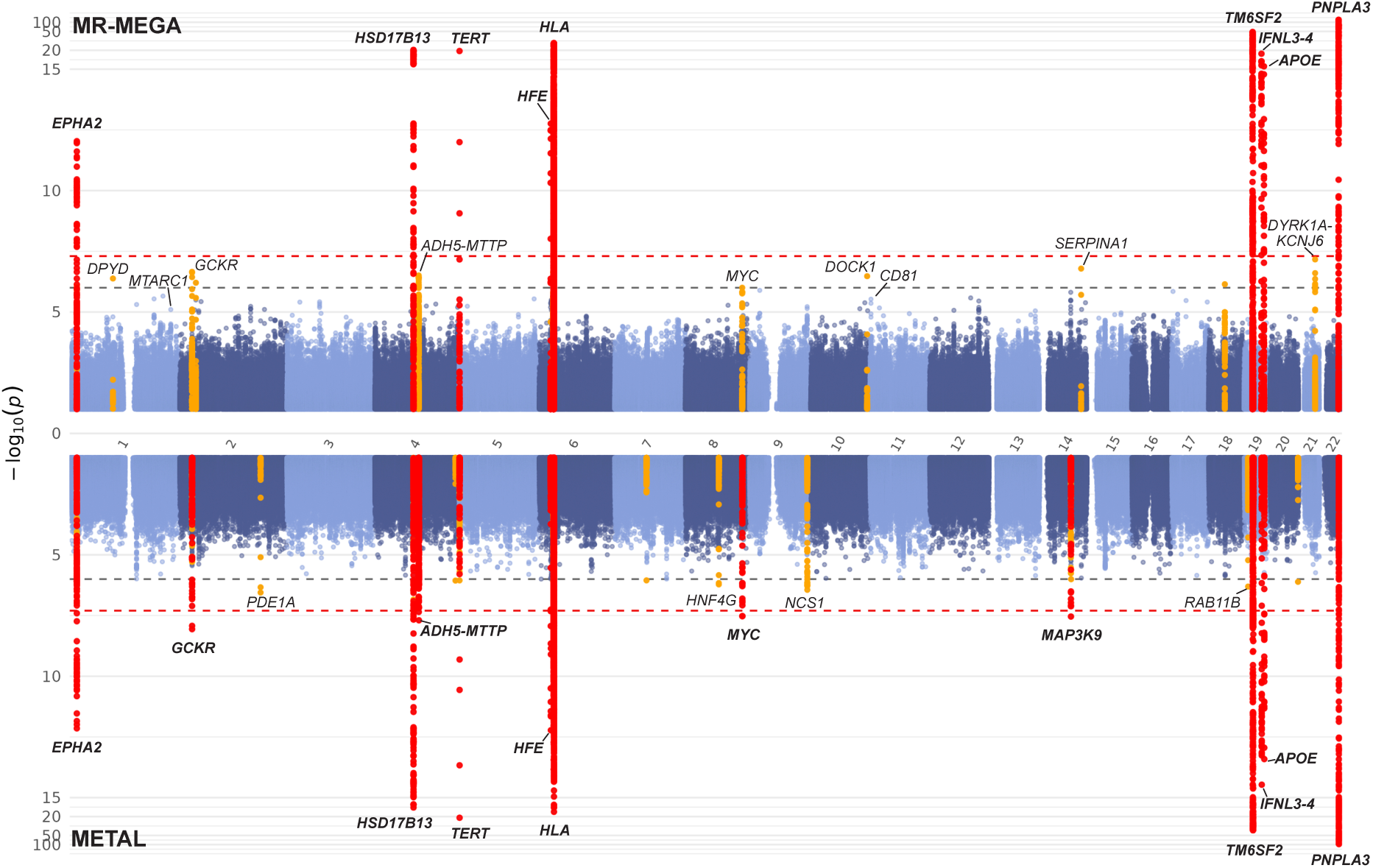
Miami plot of the meta-analysis results from 17,329 HCC cases and 2,424,298 controls. Results from MR-MEGA (top) and METAL (including all ancestries, bottom) with genome-wide significant (P < 5x10^-8^) loci in red and top suggestive loci in orange.

**Table 1.**
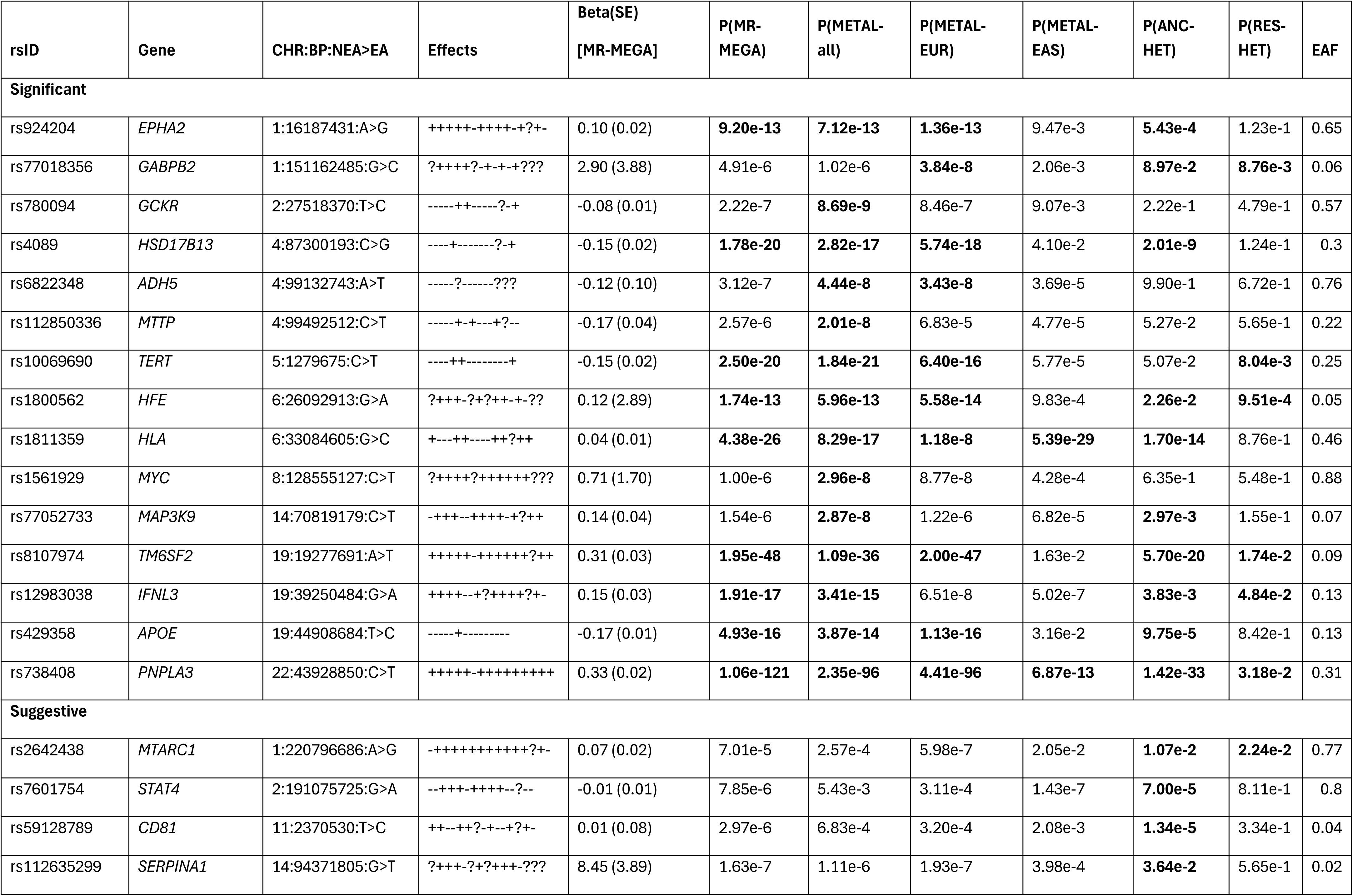
GWAS-significant loci associated with HCC from MR-MEGA and METAL meta-analysis models. Effects indicates the direction of effect on meta-analysis for each study where + represents a positive beta and – a negative beta, andwhere this variant was not included in the dataset. Studies included under ‘Effects’ in order (from left to right): Biobank Japan, Hassan et al., DeCode, Finngen, Intermountain, KoGES, PLCO, Trepo et al., UKBB, MVP (EUR), MVP (AFR), MVP (AMR), All of Us, TPMI, CKBB. CHR, chromosome; BP, base pair; NEA, non-effect allele; EA, effect allele. BETA(SE), allelic effect in log odds ratio from MR-MEGA analysis; P(MR-MEGA): two-sided P value of association from MR-MEGA (chi-squared test with df = 4); P(METAL-all), two-sided P value of association from sample-size weighted analysis using METAL with all ancestries; P(METAL-EUR/EAS), two-sided P value of association from sample-size weighted analysis using METAL with European (EUR) or East Asian (EAS) ancestries; P(ANC-HET), P value for the two-sided ancestral heterogeneity test (chi-squared test with df = 3); P(RES-HET): P value for the two-sided residual heterogeneity test (chi-squared test with df = 3); EAF, effect allele frequency. Bolded are all significant P values (P < 5x10^−8^ for the two-sided association tests, P < 0.05 for the heterogeneity tests).

Three novel GWAS-significant loci (*GCKR*, *ADH-1B/-4/-5, and MTTP*), are well established risk genes for liver disease. *GCKR* (lead SNP rs780094) is associated with MASLD(10) and plasma protein levels of *GCKR* are associated with HCC on Mendelian randomisation analyses(7), though not with cirrhosis (Supplementary Table 5). The *ADH-1B/-4/-5* locus (lead SNP rs6822348) is a genome-wide significant locus for cirrhosis(12), confirmed through colocalization analysis (probability of shared causal variant (PP.H4.abf) = 0.90, Supplementary Table 5), which may mediated through effect on alcohol intake (Figure 3A). The causal gene in this region remains unclear as, whilst typically annotated as *ADH1B* or *ADH4*, rs6822348 has a liver-specific eQTL for *ADH5* (-0.17 normalised effect size, p= 8.1x10^-8^, from GTEx). It should be noted that Vujkovic *et al.* identified a different, coding lead variant in ADH1B (rs1229984 encoding p.His48Arg) to be associated with cirrhosis, which is not in LD with rs6822348 (R^2^ < 0.1), suggesting there may be more than one effect in this locus.

**Fig 3.**
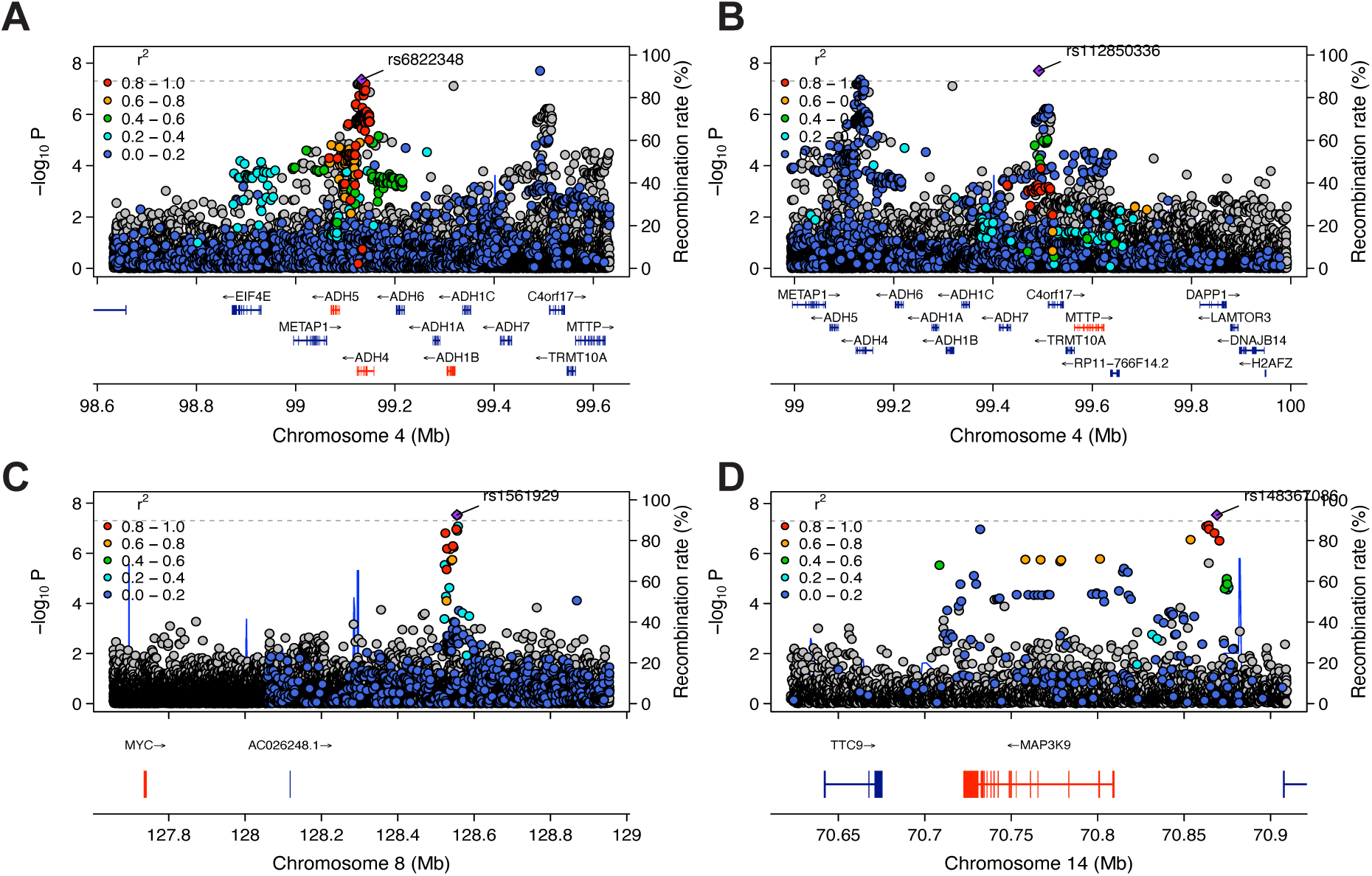
Region plots of novel genome-wide significant associations. LocusZoom plots of genome-wide significant (P < 5x10^-8^) loci demonstrating individual study effects for: *ADH5* (A), *MTTP* (B), *MYC* (C), and *MAP3K9* (D).

*MTTP* (lead SNP rs112850336) is central to hepatic triglyceride metabolism which is genome-wide significant for MASLD(10). *MTTP* is less than 1MB upstream of *ADH5*, but these genes were demonstrated to have independent effects on liver fat in the MASLD GWAS(10). Here, we observed separate peaks and the lead variants (rs6822348 near *ADH5* and rs112850336 near *MTTP*) were not in LD (R^2^=0.1, Figure 3 A-B).

SNP rs1561929 demonstrated consistent effects across all cohorts where present. The closest protein coding gene is *MYC* (Fig 3C), which is amplified or has gain of function mutations in 227/373 (61%) HCC cases based on The Cancer Genome Atlas Program (TCGA) data. *MYC* is a common essential gene (including liver cancer cell lines) in DepMap(13) and has a probability of being loss-of-function intolerant (pLI) score of 1.0(14).

Variants in *MAP3K9* (lead SNP rs77052733) were also positively associated with HCC (Fig 3D). 110/373 (29.5%) of HCC cases from TCGA have heterozygous deletion of *MAP3K9*.

Lastly, we identified a novel GWAS-significant locus for EUR ancestry at near *GABPB2* (lead SNP rs77018356, SupFig 3 A-B). There was no evidence of colocalization with variants causing cirrhosis for *MYC*, *MAP3K9*, *EPHA2*, or *GABPB2* loci.

Our analyses also replicated loci in *SERPINA1*, *CD81*, *STAT4*, *MTARC1* at least suggestive significance (P <5x10^-6^), in addition to 57 additional loci (Supplementary Table 4, SupFig 3 C-D). Prioritisation of these loci based on pLI found 13 genes with pLI 0.99-1, including *HFN4G*, *MEN1* (SupFig 3 E-F), and *BRINP3*, which may be worthy of further study.

### Cirrhosis-independent effects

Most signals found are in loci previously linked to development of cirrhosis, therefore we ran a sensitivity analysis in two cohorts with cirrhotic controls (Trepo *et al.* and Hassan *et al.*).

We found consistent effects in *APOE*, *EPHA2*, *GABPB2*, *HSD17B13*, *MAP3K9*, *PNPLA3*, *TERT*, and *TM6SF2*, suggesting that these loci increase risk of HCC even compared to cirrhotic controls (Supplementary Table 6). There were no variants included in both studies suitable as a proxy for rs1800562 in HFE. The effects of *ADH5*, *GCKR*, HLA region, *IFNL3*, and *MYC* were attenuated, suggesting that some of their effect may be via underlying cirrhosis, though there was reduced power in this two study sensitivity analysis.

### MAMA identifies heterogenous effects across ancestries

5 loci showed homogenous effects across different ancestries (P_ANC-HET_ > 0.05), including variants in or near *TERT*, *GCKR*, *ADH5, MTTP*, and *MYC* (Figure 4A-C, Table 1). For 6 loci, (*IFNL3-IFNL4*, *EPHA2*, *MAP3K9*, *HSD17B13*, *APOE*, and *HLA* loci) we found significant ancestral heterogeneity (P_ANC-HET_ < 0.05) without residual heterogeneity (P_RES-HET_ > 0.05), suggesting that the results are due to population structural differences instead of other confounders (Figure 4D). Only three well-established HCC loci (*PNPLA3*, *TM6SF2*, *HFE*) had both significant ancestral heterogeneity and residual heterogeneity.

**Fig 4.**
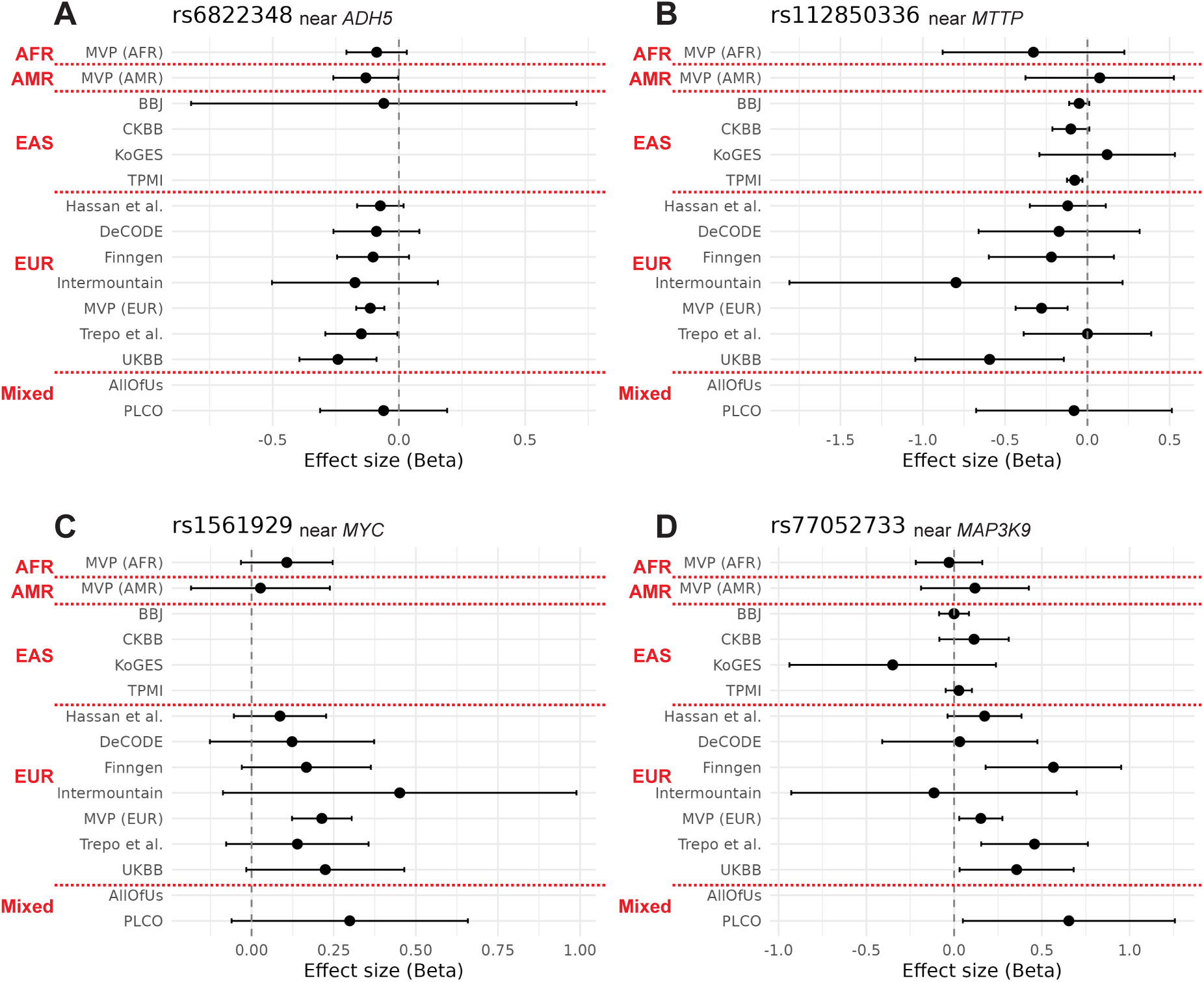
Forest plots of novel genome-wide significant associations. Forest plots of genome-wide significant (P < 5x10^-8^) loci (top) with forest plots demonstrating individual study effects (below) for: *ADH5* (A), *MTTP* (C), *MYC* (C), and *MAP3K9* (D). BBJ, BioBank Japan; CKBB, China Kadoorie BioBank; KoGES; Korean BioBank; MVP, Million Veterans Programme; PLCO, Prostate, Lung, Colorectal and Ovarian (PLCO) Cancer Screening Trial; TPMI, Taiwan Precision Medicine Initiative; and UKBB, UK BioBank. Annotated with genetic ancestry: AFR, African; AMR, Admixed American; EAS, East Asian; EUR, European.

Of note, the *HLA* locus (lead SNP rs1811359, P_ANC-HET_ = 1.7x10^-14^) has strongly positive effect estimates in East Asian cohorts, whilst negative effect estimates in European ancestry cohorts (SupFig 3G). The AFR and AMR cohorts demonstrated neutral effects at the *HLA* locus.

Five loci had significant effects in European ancestry cohorts and MAMA but no effect in EAS cohorts. The *TM6SF2* locus (lead SNP rs8107974, P_ANC-HET_ = 5.7x10^-20^) has strongly positive in AMR and EUR ancestry cohorts whilst no effect in AFR and EAS cohorts (P_(METAL-EAS)_ = .016, SupFig 3H). Similarly, the *EPHA2*, *APOE*, *HFE*, and *HSD17B13* loci had no significant effects in EAS cohorts (P > 1x10^-5^) but were directionally consistent with genome-wide significant effects in EUR cohorts. These observations likely reflect the influence of the variants on cirrhosis, as the primary mediator of HCC, in their respective ancestries.

### Fine mapping identifies 11 missense variants

MR-MEGA was used to generate posterior probabilities for fine mapping, which found 28 significant or suggestive loci that had fewer than 5 variants in the 95% credible set. Within this, we found putative causal coding variants in 11 loci (Table 2 and Supplementary Table 7). This includes the well-established missense mutations in PNPLA3, TM6SF2, HFE, APOE, SERPINA1, and MTARC1. In addition, identified GCKR p.Leu446Pro as the likely causal variant; this missense mutation is strongly associated with insulin resistance and hepatic steatosis.

**Table 2.** Coding variants identified from fine mapping. Posterior probabilities were calculated from Bayes scores from MR-MEGA meta-analysis results for all significant and suggestive loci. Loci were filtered for those with fewer than 5 variants per 95% credible set. Ensembl Variant Effect Predictor was used to identify common coding variants from those credible sets. CADD, Combined Annotation Dependent Depletion score; CHR, chromosome.

| CHR:Start:End | Variant | P-value | Gene | Variant | CADD |
| --- | --- | --- | --- | --- | --- |
| 22:43927717:44014293 | 22:43928847:C>G | 1.06e-121 (MR-MEGA) | PNPLA3 | p.Ile148Met | 13.4 |
| 19:18952750:20375946 | 19:19268740:C>T | 1.95e-48 (MR-MEGA) | TM6SF2 | p.Glu167Lys | 24.6 |
| 19:44883210:44925202 | 19:44908684:T>C | 4.93e-16 (MR-MEGA) | APOE | p.Cys130Arg | 16.6 |
| 6:25526091:27552168 | 6:26092913:G>A | 1.74e-13 (MR-MEGA) | HFE | p.Cys176Tyr | 27 |
| 2:27375230:27529596 | 2:27508073:T>C | 8.69e-9 (METAL-all) | GCKR | p.Leu446Pro | 10.99 |
| 10:127419693:127419693 | 10:127419693:G>A | 3.35e-7 (MR-MEGA) | DOCK1 | p.Glu1553Lys | 23.6 |
| 19:8359282:8386830 | 19:8369239:G>C | 4.93e-7 (MR-MEGA) | ANGPTL4 | p.Glu190Gln | 19.01 |
| 1:220796686:220800419 | 1:220796686:A>G | 5.98e-7 (METAL-EUR) | MTARC1 | p.Thr165Pro | 18.3 |
| 6:31354452:31359387 | 6:31355519:A>G | 4.38e-26 (MR-MEGA) | HLA-B | p.Gly231= | - |
| 14:94206394:94378610 | 14:94378610:C>T | 1.63e-7 (MR-MEGA) | SERPINA1 | p.Glu366Gln | 24.2 |
| 11:64800220:64813774 | 11:64805130:G>A | 2.25e-6 (MR-MEGA) | MEN1 | p.Asp418Glu | 19.25 |

Across suggestive loci, we found MEN1 p.Asp418Glu and ANGPTL4 p.Glu190Gln missense variants. *MEN1* is a tumour suppressor gene, where heterozygous loss of function mutations cause multiple endocrine neoplasia type 1. *ANGPTL4* is a circulating protein that regulates angiogenesis and inactivation of lipoprotein lipase, therefore influences serum lipid levels(15,16).

### *DHRS1* is associated with HCC on transcriptome-wide analysis

We used transcriptome-wide eQTL data from control liver and HCC to perform a transcriptome-wide association analysis (TWAS) using FUSION (Supplementary Table 9). This replicated the above observations for *EPHA2* and *TM6SF2*. In addition, we found a novel TWAS signal in *DHRS1* (lead GWAS-SNP rs2295308, P-adj_TWAS_ = 6.3x10^-5^) based on MR-MEGA meta-analysis results and HCC-specific eQTL (P-_eQTL_ = 5.1x10^-5^). *DHRS1* is thought to have oxioreductase activity(17) though its function in the liver or malignancy has not yet been characterised.

We subsequently performed a regulome-wide association study (RWAS) to identify regions where germline variation in chromatin accessibility influence risk of HCC (Supplementary Table 9). Here, we discovered signals for the above GWAS-significant genes *HSD17B13*, *HFE*, *TM6SF2*, and *PNPLA3*. In addition, we found a novel highly significant chromatic accessible region at chr9:96436312-96436813 (lead GWAS-SNP rs16911041, P-adj_RWAS_ = 2.7x10^-24^). The nearest protein coding genes are *ZNF367*, a transcription factor with tumour suppressor properties(18), and *HABP4*, which is implicated in familial colorectal cancer(19).

### Gene set analysis shows enrichment in liver

We ran multi-marker analysis of genomic annotation (MAGMA) for gene ontology, tissue level and single-cell expression data(20). Enriched gene ontology sets in MSigDB v7.0 were dominated by those related to antigen presentation given mapping to multiple HLA genes. (Supplementary Figure 4A and Supplementary Table 8). In addition, we observed enrichment of the β-catenin-TCF transactivating complex based on *TERT*, *MEN1*, and histone genes (P-adj=.03) and GWAS catalogue signatures related to chronic liver disease. Differentially expressed genes were preferentially expressed in liver tissue, based on GTEx data (Supplementary Figure 4B).

### HCC shares genetic architecture with chronic liver disease but not other gastrointestinal cancers

Finally, we used linkage disequilibrium score regression to quantify the similarity in genetic architecture between HCC, liver disorders, and other gastrointestinal cancers (Figure 5).

There was a strong positive genomic correlation between HCC and cirrhosis (rg=0.87, p=5.0x10^-10^), HBV (rg=0.84, p=.003), HCV (rg=0.65, p=.004), and MASLD (rg=0.50, p=6.0x10^-4^), reflecting the underlying drivers of HCC in the population. There was no significant correlation between HCC and upper GI cancer (oesophageal and gastric) or colorectal cancer. There were shared genetic determinants between HCC and T2DM (rg=0.25, p=6.8x10^-5^), whilst there was inverse association between HCC and BMI (rg=-0.17, p=6.0x10^-4^), suggesting that genetically lower BMI is linked to correlated with increased genetic risk of HCC.

## Discussion

This large GWAS meta-analysis of all-cause HCC has discovered evidence for germline involvement of several novel genes involved in HCC, most notably *MYC* and *MAP3K9*. We also found genome-wide significant germline associations for HCC in *GCKR*, *ADH5*, *MTTP* for the first time. In addition, we have dissected diverse effects across ancestries and leveraged these results to fine-map 11 missense variants. Finally, we demonstrate the importance of germline risk for cirrhosis, T2DM, and low BMI as drivers of population risk for HCC.

*MYC* is an archetypal proto-oncogene. It is a transcription factor that lies downstream of multiple cell surface signalling pathways, including the WNT-β-catenin pathway(21), which is robustly implicated in HCC development(22–24). Common variation near *MYC* confers increased susceptibility to *WNT* signalling in other malignancies(25). These results suggest a similar process may be involved in germline risk of HCC development. It is unclear whether its effect is dependent on cirrhosis or not; there was no evidence of co-localisation but it was not significant on sensitivity analysis using only cirrhotic controls.

The mitogen activated protein kinase kinase kinases (MAP3K) are a group of proteins that act downstream of cellular stress signals and growth factors receptors. They function as part of a signalling cascade that ultimately leads to pro-growth effects. *MAP3K9* primarily transduces cellular stress through to *JUN* and *GATA4*, which influences mitochondrial cytochrome c release and therefore apoptosis(26,27). Mutations in *MAP3K9* have been identified in lung cancer and melanoma(28,29), as well as in HCC via the TCGA dataset. It is not knows precisely which stress signals regular *MAP3K9* in the liver, though interactions with a variety of microRNAs have been proposed(30,31).

Prevalence of HCC vary greatly across the globe, from 1/100,000 in Morocco to 16/100,000 in Republic of Korea(32). Whilst this is driven by differences in the prevalence of chronic viral hepatitis, alcohol intake, and obesity, our analysis has uncovered ancestral heterogeneity in germline contribution to HCC risk. Specific HLA loci showed divergent effects in European and East Asian ancestries. Also, in individuals of East Asian ancestry, we observed no significant effect from *TM6SF2*, which was the most significant locus in Europeans(33).

Some these differences may reflect the underlying aetiology of HCC in these regions (i.e. viral hepatitis versus steatotic liver disease).

Genetically driven accumulation of hepatic lipid droplets has been associated with severity of almost every studied liver disease. In this analysis we found evidence for six genes (*MTTP*, *PNPLA3*, *TM6SF2*, *HSD17B13*, *GCKR*, and *APO*) that are all genome-wide risk genes for MASLD(10). Gene-environment studies have previously shown that the effect of such variants is amplified by the presence of obesity, type 2 diabetes, and excess alcohol consumption(34). The stronger effects observed in European cohorts may reflect the higher prevalence of steatotic liver disease in these populations(35). It should also be noted that non-cirrhotic HCC is well described in steatotic liver disease(36). These data underscore the importance of public health interventions to tackle HCC.

The principal strengths of this study are its size and TWAS/RWAS analyses. This study would be improved by inclusion of individual participant level data, which would allow for mediation analyses as performed by Vujkovic et al. Also, most cohorts used electronic health records for identification of cases, which has the risk of mis-classification.

Despite the inclusion of multiple ancestries, individuals of European ancestry are still over-represented and account for 49% of HCC cases in this analysis. Individuals of African descent are particularly under-represented, with only 6% of all cases and no data from the continent of Africa, which has some of the highest rates of HCC(37). We are also lacking data from the Middle East and South America, where HCC are top causes of cancer death. Addition of data from population biobank studies from these areas will strengthen future MAMA.

Most cohorts included in this analysis had healthy population controls, therefore this meta-analysis reflects the population-level drivers of HCC. Therefore, many loci are already implicated in development of underlying liver disease (e.g. *TM6SF2*). Our sensitivity analysis with cirrhosis controls and colocalisation studies demonstrated that most significant loci, including *MAP3K9*, appears to act independently of cirrhosis. Larger meta-analyses with cirrhotic controls or aetiology-specific studies may identify further novel treatment targets.

## Conclusion

HCC is driven by heterogenous and homogenous genetic factors across ancestries. Germline susceptibility to beta-catenin pathway activation influences HCC risk via *MYC*, *MEN1*, and *TERT*. MAP3K9 and *DHRS1* are novel HCC risk genes worthy of further investigation. Genes involved in hepatic lipid droplet formation, including *MTTP*, contribute to risk of HCC. There is need for further ancestral diversity in liver GWAS.

## Methods

### Study design and cohort descriptions

We used a single-stage joint meta-analysis design to maximize statistical power: (1) MR-MEGA for heterogenous effects across ancestries; and (2) sample-size weighted analysis via METAL for homogeneous effects across all ancestries as well as within European (EUR) and East Asian (EAS) ancestries. We used datasets representing four different ancestry groups: European, East Asian, Admixed American (AMR), African (AFR) as well as two cohorts of mixed ancestry (which were >70% European, Supplementary Table 1). GWAS results of Trepo et al.(4) were acquired through GWAS catalogue (GCST90092003).

Summary statistics for Hassan et al.(5) were previously reported and provided via direct collaboration with the authors. UKBB, DeCode, and Intermountain summary statistics were obtained from https://www.decode.com/summarydata/ and are reported previously(38). The FinnGen GWAS summary statistics were acquired from FinnGen Release 9 (https://www.finngen.fi/en ). Taiwan Precision Medicine Initiative (TPMI) summary statistics were acquired from https://pheweb.ibms.sinica.edu.tw/pheno/155.1 (39,40). China Kadoorie Biobank (CKBB) summary statistics were acquired from https://pheweb.ckbiobank.org./download/c22 (41). BioBank Japan (BBJ) summary statistics were acquired from GWAS Catalogue (GCST90018638)(42). Korean BioBank (KoGES) summary statistics were acquired from https://koges.leelabsg.org/ (43). Prostate, Lung, Colorectal and Ovarian (PLCO) Cancer Screening Trial summary statistics were acquired from https://exploregwas.cancer.gov/plco-atlas (44). Millions Veterans Programme (MVP) summary statistics were acquired from https://ftp.ncbi.nlm.nih.gov/dbgap/studies/phs002453/analyses/GIA/ as three separate cohorts for EUR, AMR, and AFR, and are reported previously(45). All of Us summary statistics were generated through analyses for this study(46,47).

GRCh38 were used for analysis and reported throughout. Where GRCh37 was required by specific tools (e.g. FUMA), LiftOver from MungeSumstats(48) was used for conversion.

### All of US

Inclusion criteria and genotyping of the All of Us cohort is described in detail elsewhere(47). In brief, All of Us is a population-level dataset of approximately 350,000 individuals from the United States of America. All of Us data was accessed using Research Project ID 65834.

Individuals were included in the current analysis if there was available data on age, sex, and presence or absence of HCC. ICD diagnostic codes (C22.0-ICD03, 155.0-ICD9, and C22.8-ICD10) were used to identify HCC cases. GWAS was run using Hail(49) adjusted for age, sex, and the first 9 principal components (PC) of genetic ancestry.

### Multi-ancestry meta-analysis (MAMA)

We performed MAMA of GWAS results using MR-MEGA(50) and METAL(51). MR-MEGA accounts for differences in population structure through meta-regression, where axes of genetic variation generate covariates for inclusion in the meta-analysis to. Four PC were included in the analysis. MR-MEGA has reduced power to detect associations for variants with homogeneous effects across populations. It is therefore recommended to run MR-MEGA in conjunction with another methods, we therefore used sample size weighted METAL (as has been reported in two recent meta-analyses(6,7)) to detect homogenous allelic effects.

Prior to meta-analysis, all datasets were harmonised to genome build hg38 using MungeSumstats(48) and R 4.5.0(52) and included in NCBI GRCh38 (2013-12-17) reference panel. Sex-chromosome data were not available for all cohorts, therefore only autosomal variants were kept in the results. In total 11,122,532 variants were included in MR-MEGA analyses, as this method has a cohort-number requirement that varies based on the number of axes of variation. Number of variants included in METAL analyses were: 33,182,383 for all cohorts, 22,029,853 for EUR, and 7,735,436 for EAS. Genomic inflations were measured for all cohorts and the meta-analyses. Genomic control was applied to meta-analysis using METAL. All inflation was nominal and below 1.02 for all cohorts and results. GWAS-significance was set at P < 5x10^-8^ and suggestive threshold set at P < 1x10^-6^. For variants to be considered GWAS-significant in all ancestry analysis using METAL, they must also reach at least suggestive significance with MR-MEGA.

We identified genomic risk loci within our meta-analysis results using Functional Mapping and Annotation (FUMA) v1.8.1(53,54). FUMA identifies independent significant SNPs in the GWAS results by clumping significant variants (r^2^ threshold < 0.6), then defines the locus by merging linkage disequilibrium (LD) blocks of all independent significant SNPs within 250 kb. Start and end of a locus is a region that encompasses all independent significant SNPs (r^2^ ≥ 0.6). Further clumping of the independent significant variants within the locus (r^2^ ≥ 0.1) defines the lead SNP.

All significant SNPs were compared to the known HCC risk variants reported in previous analyses, including the pre-print by Vujkovic *et al*(7). Forest plots and Manhattan plots were generated using R 4.5.0. QQ plots were generated by FUMA.

#### Colocalisation

To determine whether variants were shared between HCC and cirrhosis, we performed colocalization analysis using summary statistics from the largest available GWAS of cirrhosis(12). Summary statistics were standardised using MungeSumstats and converted into variant call format using gwas2vcf for analysis using gwasglue(55). Evidence of colocalization due to a common causal variant was defined by PP.H4 ≥0.8.

#### Fine-mapping

Fine-mapping was performed using MR-MEGA, which derives a natural log of Bayes factor (lnBF) per SNP in favour of association, as previously described(56). All significant and suggestive SNPs were included for fine-mapping. Posterior probabilities (PPs) of driving the association signal at each locus were calculated from the Bayes factor, as previously described(56). Credible sets of fewer than 5 SNPs with sum PP greater than 0.95 were accepted as putative causal variants. Fine mapped variants were run through Variant Effect Predictor (VEP) to determine functional consequences(57).

#### MAGMA: gene ontology and tissue-enrichment

Results were analysed using MAGMA(54) to identify gene ontology term enrichment and gene expression data from tissues in GTEx v8, this was performed via the GENE2FUNC function of FUMA with default parameters(57). Mapped genes were analyzed using gene set analysis for ontology terms from MSigDB v7 and gene-property analysis for tissue specificity. Results were adjusted for multiple tests using Bonferroni FDR correction with the alpha of 0.05. The significant ontology terms were filtered for terms that share the same signals. Tissue level enrichment analysis was done using the pre-processed GTEx gene expression dataset provided by FUMA investigators.

#### Transcriptome-wide and regulome-wide association studies

TWAS and RWAS were performed using FUSION(58), which uses tissue- (or cancer-) specific eQTLs to determine the effects of germline variation on transcriptome profile (TWAS) or chromatic accessibility (RWAS)(59). TWAS was performed using (i) GTEx v8 liver pre-computed weights, and (ii) HCC pre-computed weights (‘LIHC’), as provided by the FUSION investigators, with 1000Genomes data reference panels. RWAS was performed known chromatin accessibility profiles for HCC (‘LIHC’). All results underwent Benjamini–Hochberg FDR correction with the alpha of 0.05.

#### LDSC regression analysis across traits

We performed linkage disequilibrium score regression (LDSC) analysis to compare the similarity in genetic architecture across traits. Summary statistics for the largest publicly available GWAS were obtained for: cirrhosis (GCST90319878)(12), liver fat (GCST90016673)(60), alanine transaminase (ALT, GCST90018943)(42), hepatitis B viral infection (HBV, GCST90479751)(59), hepatitis C virus infection (HCV, GCST90018805)(60), body mass index (BMI, GCST009004)(61), type 2 diabetes mellitus (T2DM, GCST90444202)(62), upper gastrointestinal (GI) cancer (gastric and oesophageal, GCST90011807)(63), and colorectal cancer (GCST90129505)(64). Summary statistics were formatted and converted to GRCh37 using MungeSumstats then aligned to 1000 Genomes reference panel. We then ran pairwise regression (--rg) comparisons between all traits.

## Data availability

Raw data for Hassan *et al.* is available via dbGaP phs001744.v1.p1. All other summary statistics are publicly available via the above links. Full summary statistics from this analysis will be made available via GWAS catalogue upon publication. All code used in analysis is available at: https://github.com/jmann01/HCC-gwas

## Acknowledgements

We thank all the participants, investigators, and funders who contributed to the studies included in this meta-analysis.

## Supplementary data

**Supplementary Table 1.** Summary of characteristics of included cohorts.

**Supplementary Table 2.** LDSC genetic covariance intercept (gcov_int) between cohorts with EAS and within EUR ancestries with their 95% confidence intervals (CI).

**Supplementary Table 3.** Previously reported significant/suggestive loci for HCC from any cause annotated with results from MR-MEGA meta-analysis from this study.

**Supplementary Table 4.** Suggestive and significant loci associated with HCC from MR-MEGA and METAL meta-analysis models. CHR, chromosome; BP, base pair; NEA, non-effect allele; EA, effect allele. BETA(SE), allelic effect in log odds ratio from MR-MEGA analysis; BETA(SE)_METAL, allelic effect in log odds ratio from METAL (all ancestries) analysis;P(MR-MEGA): two-sided P value of association from MR-MEGA (chi-squared test with df = 4); P(METAL-all), two-sided P value of association from sample-size weighted analysis using METAL with all ancestries; P(METAL-EUR/EAS), two-sided P value of association from sample-size weighted analysis using METAL with European (EUR) or East Asian (EAS) ancestries; P(ANC-HET), P value for the two-sided ancestral heterogeneity test (chi-squared test with df = 3); P(RES-HET): P value for the two-sided residual heterogeneity test (chi-squared test with df = 3); EAF, effect allele frequency.

**Supplementary Table 5.** Results from colocalisation analyses. Where: PP.H0.abf = Probability neither HCC or cirrhosis are associated; PP.H1.abf = Probability only HCC is associated; PP.H2.abf = Probability only cirrhosis is associated; PP.H3.abf = Probability both traits are associated, but with different causal variants; PP.H4.abf = Probability both traits are associated and share the same causal variant. nsnps = number of SNPs included in coloc analysis.

**Supplementary Table 6.** Results from sensitivity analysis in cirrhotic controls: meta-analysis of Trepo et al. and Hassan et al. using METAL with sample size weighting, with focused analysis of previously identified significant and suggestive loci (from any previous analysis). P-values underwent Bonferroni adjustment for multiple tests.

**Supplementary Table 7.** Fine-mapping results. Posterior probabilities were calculated from Bayes scores from MR-MEGA meta-analysis results for all significant and suggestive loci.

Loci were filtered for those with fewer than 5 variants per 95% credible set. Ensembl Variant Effect Predictor was used to annotate variants within the credible sets, keeping coding variants and those with high CADD scores. CADD, Combined Annotation Dependent Depletion score; CHR, chromosome; nGWAS-SNPs, number of SNPs with the locus that met significant/suggestive thresholds.

**Supplementary Table 8.** Pathway analysis using MAGMA based on mapped genes from FUMA analysis. P-values were Bonferroni adjusted and only significantly enriched pathways are shown.

**Supplementary Table 9.** Transcriptomie-wide analysis (TWAS) results. FUSION was used to perform TWAS using normal liver tissue and hepatocellular carcinoma (HCC) eQTLs.

TWAS p-values were adjusted for multiple testing with Benjamini–Hochberg correction with the alpha of 0.05.

**Supplementary Table 10.** Regulome-wide analysis (RWAS) results. FUSION was used to perform RWAS using chromatin accessibility profiles for hepatocellular carcinoma (HCC) eQTLs. RWAS p-values were adjusted for multiple testing with Benjamini–Hochberg correction with the alpha of 0.05.

**Supplementary Table 11.** Summary of linkage disequilibrium score regression (LDSC) correlation analyses between hepatocellular carcinoma (HCC) and related traits. BMI, body mass index; HBV, hepatitis B virus; HCV; hepatitis C virus; MASLD, metabolic dysfunction-associated steatotic liver disease; T2DM, type 2 diabetes mellitus; upper GI (gastrointestinal) cancer, oesophageal and gastric cancer.

---

**Supplementary Figure 1.**
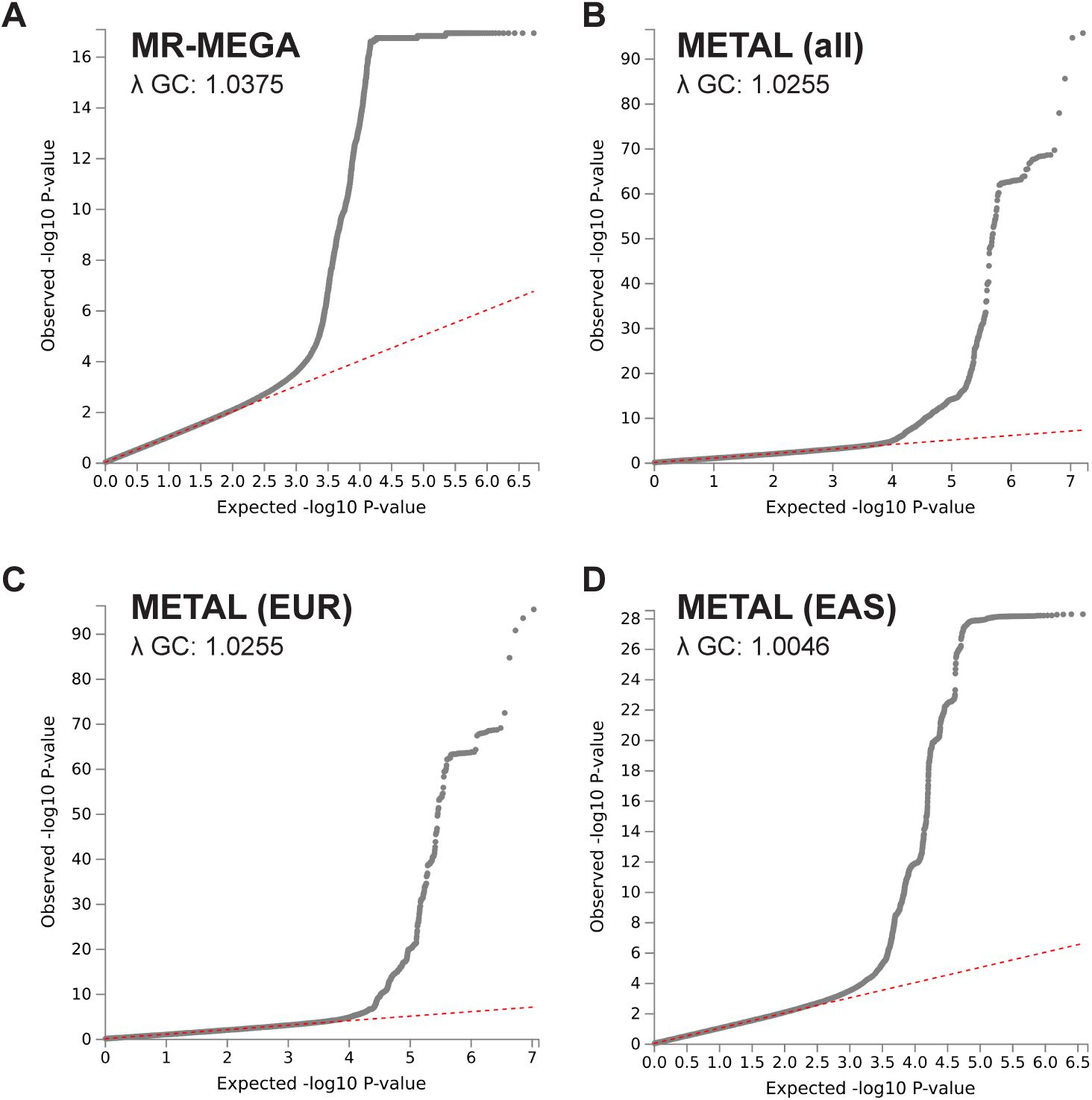
Quantile-quantile (QQ) plots for all four meta-analysis results, annotated with genomic inflation scores (lambda GC) derived from linkage disequilibrium score regression (LDSC) analysis using ldsc.

**Supplementary Figure 2.**
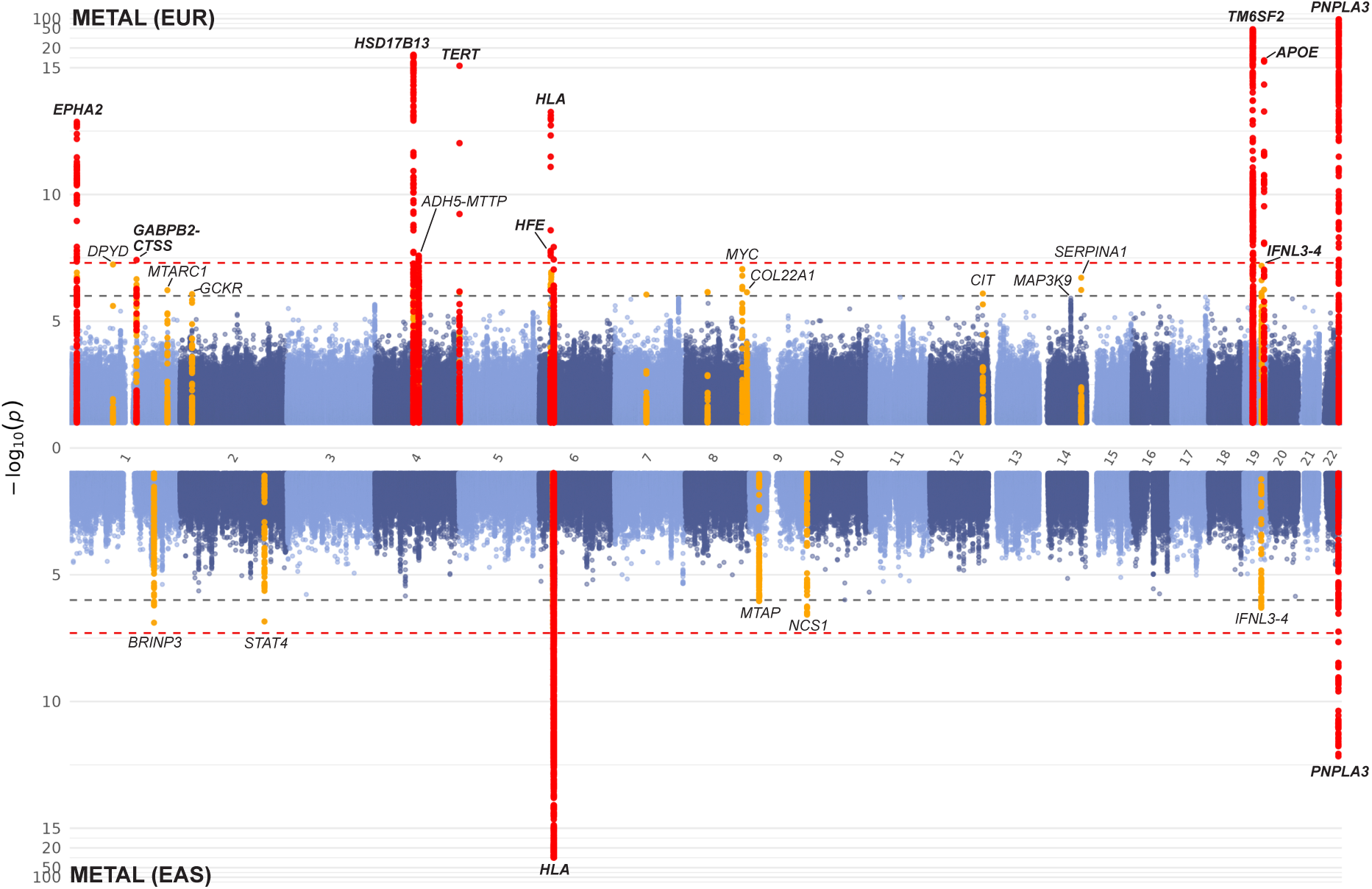
Miami plot of single-ancestry meta-analysis results. Results from METAL European meta-analysis (8,216 cases and 1,558,609 controls in 7 cohorts, top) and METAL East Asian meta-analysis (7,355 cases and 629,454 controls in 4 cohorts, bottom) with genome-wide significant (P < 5x10^-8^) loci in red and top suggestive loci in orange.

**Supplementary Figure 3.**
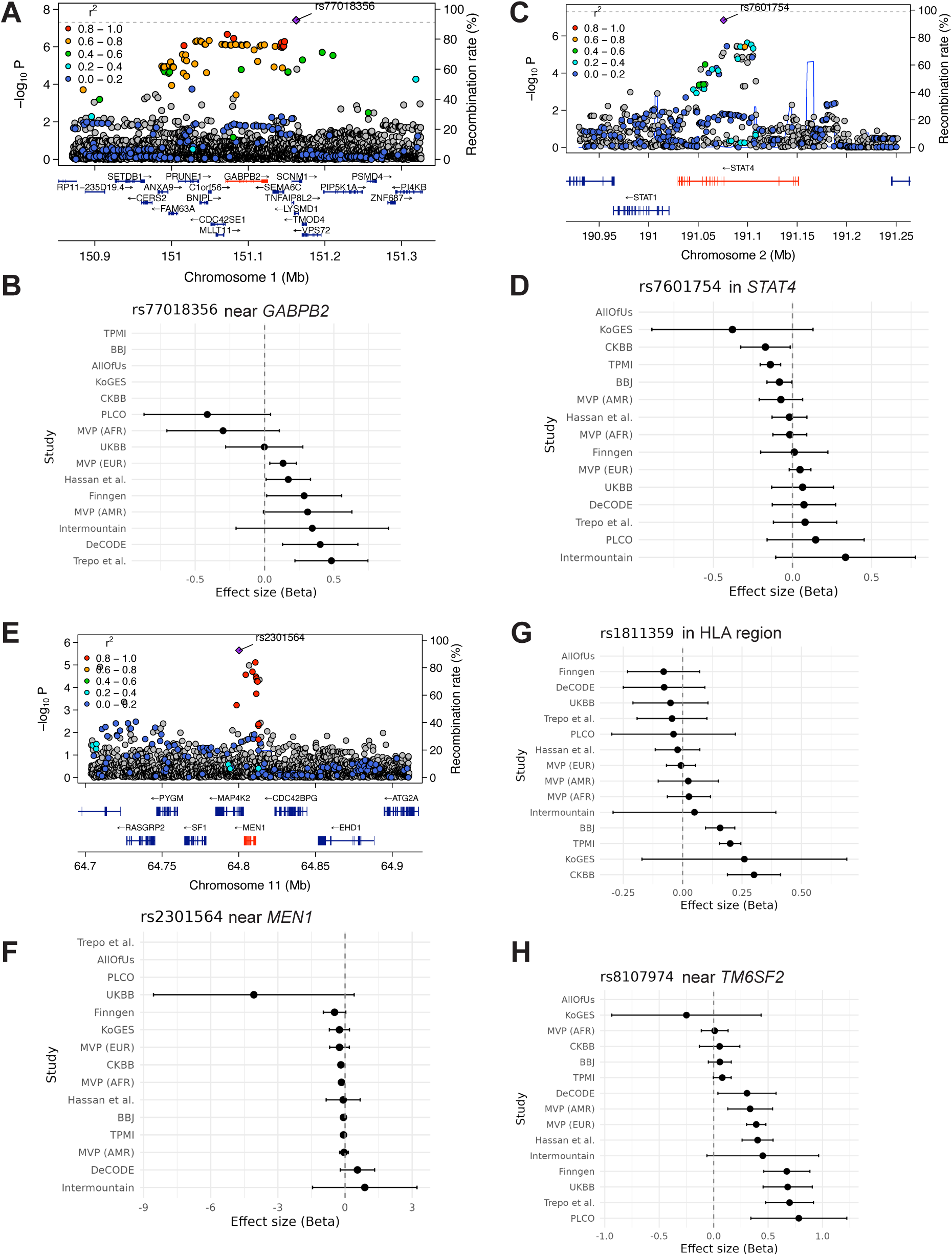
Region plots of novel suggestive associations. LocusZoom plots of genome-wide significant (P < 5x10^-8^) loci (top) with forest plots demonstrating individual study effects (below) for: *GABPB2* (A-B), *STAT4* (C-D), and *MEN1* (E-F). BBJ, BioBank Japan; CKBB, China Kadoorie BioBank; KoGES; Korean BioBank; MVP, Million Veterans Programme; PLCO, Prostate, Lung, Colorectal and Ovarian (PLCO) Cancer Screening Trial; TPMI, Thaiwan Precision Medicine Initiative; and UKBB, UK BioBank.

**Supplementary Figure 4.**
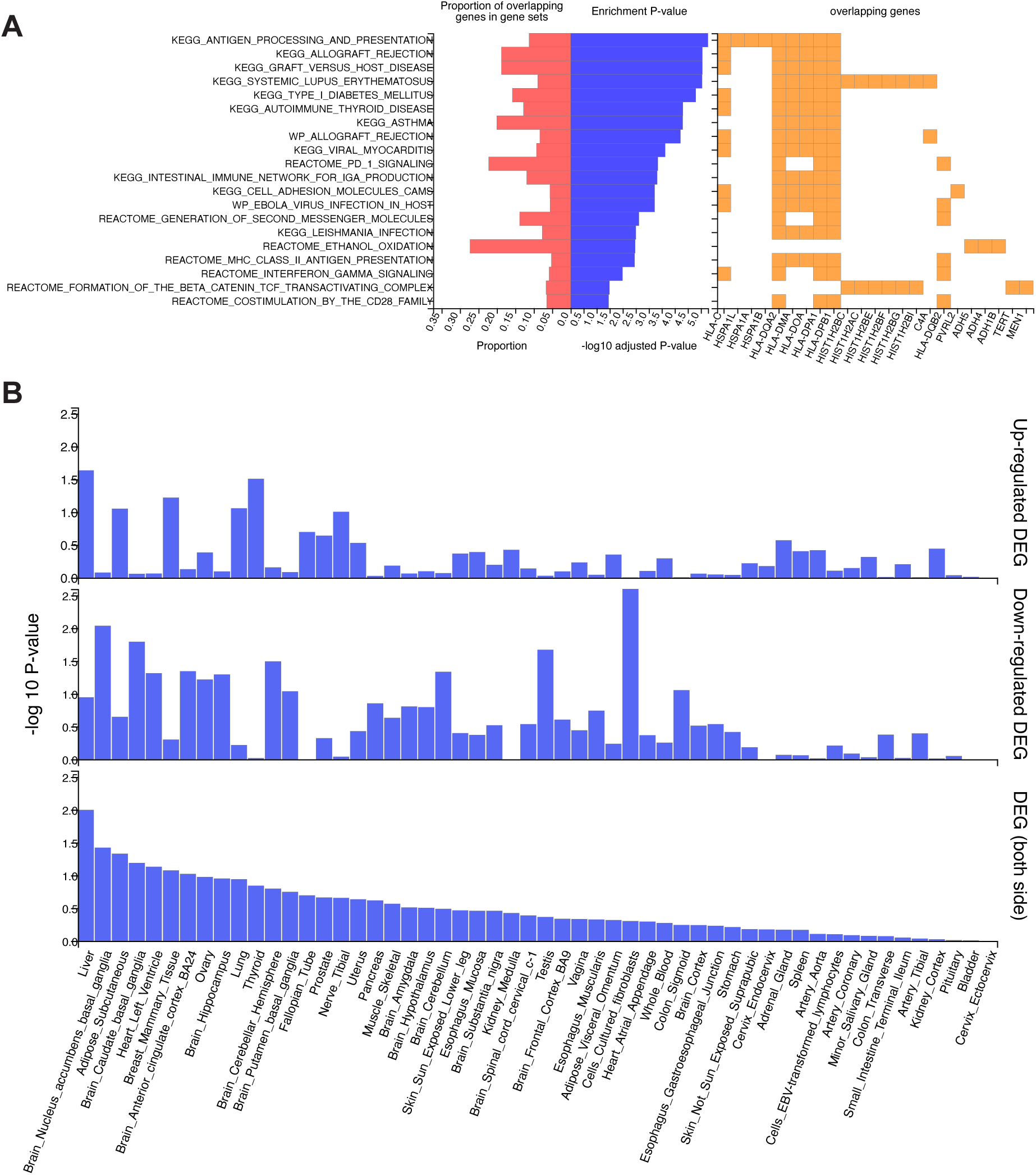
Pathway analysis results. MAGMA was used to perform gene set analysis using mapped genes from FUMA based on the METAL-all cohorts analysis. Significantly enriched canonical pathways (A) are based on Bonferroni-adjusted P-value <.05. Tissue enrichment was determined using GTEx v8 data based on up-regulation or down-regulation in the GWAS results (B).

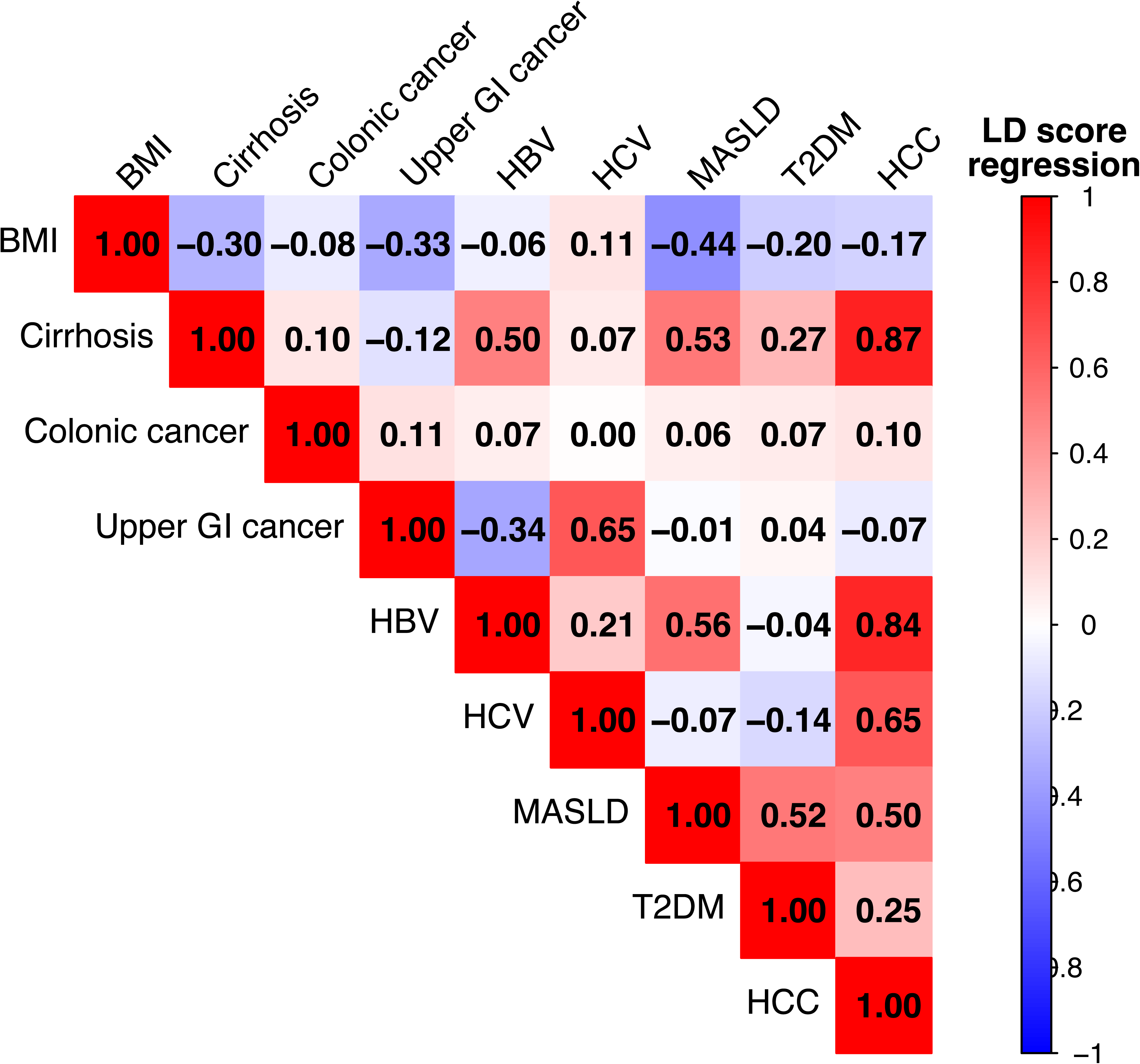

## References

1. Bray F, Laversanne M, Sung H, Ferlay J, Siegel RL, Soerjomataram I, et al. Global cancer statistics 2022: GLOBOCAN estimates of incidence and mortality worldwide for 36 cancers in 185 countries. CA Cancer J. Clin. 2024;74:229–263.

2. Singal AG, Kanwal F, Llovet J. Global trends in hepatocellular carcinoma epidemiology: implications for screening, prevention and therapy. Nat. Rev. Clin. Oncol. 2023;20:864–884.

3. Llovet JM, Pinyol R, Kelley RK, El-Khoueiry A, Reeves HL, Wang XW, et al. Molecular pathogenesis and systemic therapies for hepatocellular carcinoma. Nat. Cancer. 2022;3:386–401.

4. Trépo E, Caruso S, Yang J, Imbeaud S, Couchy G, Bayard Q, et al. Common genetic variation in alcohol-related hepatocellular carcinoma: a case-control genome-wide association study. Lancet Oncol. 2022;23:161–171.

5. Hassan MM, Li D, Han Y, Byun J, Hatia RI, Long E, et al. Genome-wide association study identifies high-impact susceptibility loci for HCC in North America. Hepatology. 2024;80:87–101.

6. Ghouse J, Gellert-Kristensen H, O’Rourke CJ, Seidelin A-S, Thorleifsson G, Sveinbjörnsson G, et al. Genome-wide meta-analysis identifies nine loci associated with higher risk of hepatocellular carcinoma development. JHEP Rep. 2025;7:101485.

7. Vujkovic M, Kaplan DE, Ghouse J, Loza B-L, Brancale J, Lewis A, et al. Germline variants influence chronic liver disease progression through distinct pathways [Internet]. medRxiv. 2025 [cited 2025 Oct 6];Available from: 10.1101/2025.09.16.25335186

8. Verweij N, Haas ME, Nielsen JB, Sosina OA, Kim M, Akbari P, et al. Germline Mutations in CIDEB and Protection against Liver Disease. N. Engl. J. Med. 2022;387:332–344.

9. Emdin CA, Haas ME, Khera AV, Aragam K, Chaffin M, Klarin D, et al. A missense variant in Mitochondrial Amidoxime Reducing Component 1 gene and protection against liver disease. PLoS Genet. 2020;16:e1008629.

10. Chen Y, Du X, Kuppa A, Feitosa MF, Bielak LF, O’Connell JR, et al. Genome-wide association meta-analysis identifies 17 loci associated with nonalcoholic fatty liver disease. Nat. Genet. 2023;55:1640–1650.

11. Buch S, Innes H, Lutz P, Nischalke H, Marquardt J, Fischer J, et al. Genetic variation in TERT modifies the risk of hepatocellular carcinoma in alcohol-related cirrhosis: results from a genome-wide case-control study. Gut. 2022;72:381–391.

12. Ghouse J, Sveinbjörnsson G, Vujkovic M, Seidelin A-S, Gellert-Kristensen H, Ahlberg G, et al. Integrative common and rare variant analyses provide insights into the genetic architecture of liver cirrhosis. Nat. Genet. 2024;56:827–837.

13. DepMap: The Cancer Dependency Map Project at Broad Institute [Internet]. [cited 2025 Nov 7];Available from: https://depmap.org/portal/

14. Chen S, Francioli LC, Goodrich JK, Collins RL, Kanai M, Wang Q, et al. A genomic mutational constraint map using variation in 76,156 human genomes. Nature. 2024;625:92–100.

15. Gealekman O, Burkart A, Chouinard M, Nicoloro SM, Straubhaar J, Corvera S. Enhanced angiogenesis in obesity and in response to PPARgamma activators through adipocyte VEGF and ANGPTL4 production. Am. J. Physiol. Endocrinol. Metab. 2008;295:E1056–64.

16. Mysling S, Kristensen KK, Larsson M, Kovrov O, Bensadouen A, Jørgensen TJ, et al. The angiopoietin-like protein ANGPTL4 catalyzes unfolding of the hydrolase domain in lipoprotein lipase and the endothelial membrane protein GPIHBP1 counteracts this unfolding. Elife [Internet]. 2016 [cited 2025 Nov 28];5. Available from: 10.7554/eLife.20958

17. Zemanová L, Navrátilová H, Andrýs R, Šperková K, Andrejs J, Kozáková K, et al. Initial characterization of human DHRS1 (SDR19C1), a member of the short-chain dehydrogenase/reductase superfamily. J. Steroid Biochem. Mol. Biol. 2019;185:80–89.

18. Jain M, Zhang L, Boufraqech M, Liu-Chittenden Y, Bussey K, Demeure MJ, et al. ZNF367 inhibits cancer progression and is targeted by miR-195. PLoS One. 2014;9:e101423.

19. Gray-McGuire C, Guda K, Adrianto I, Lin CP, Natale L, Potter JD, et al. Confirmation of linkage to and localization of familial colon cancer risk haplotype on chromosome 9q22. Cancer Res. 2010;70:5409–5418.

20. de Leeuw CA, Mooij JM, Heskes T, Posthuma D. MAGMA: generalized gene-set analysis of GWAS data. PLoS Comput. Biol. 2015;11:e1004219.

21. Dang CV. MYC on the path to cancer. Cell. 2012;149:22–35.

22. Calvisi DF, Conner EA, Ladu S, Lemmer ER, Factor VM, Thorgeirsson SS. Activation of the canonical Wnt/beta-catenin pathway confers growth advantages in c-Myc/E2F1 transgenic mouse model of liver cancer. J. Hepatol. 2005;42:842–849.

23. Kim M, Lee HC, Tsedensodnom O, Hartley R, Lim Y-S, Yu E, et al. Functional interaction between Wnt3 and Frizzled-7 leads to activation of the Wnt/beta-catenin signaling pathway in hepatocellular carcinoma cells. J. Hepatol. 2008;48:780–791.

24. Bisso A, Filipuzzi M, Gamarra Figueroa GP, Brumana G, Biagioni F, Doni M, et al. Cooperation Between MYC and β-Catenin in Liver Tumorigenesis Requires Yap/Taz. Hepatology. 2020;72:1430–1443.

25. Tuupanen S, Turunen M, Lehtonen R, Hallikas O, Vanharanta S, Kivioja T, et al. The common colorectal cancer predisposition SNP rs6983267 at chromosome 8q24 confers potential to enhanced Wnt signaling. Nature genetics [Internet]. 2009 [cited 2025 Nov 28];41. Available from: 10.1038/ng.406

26. Durkin JT, Holskin BP, Kopec KK, Reed MS, Spais CM, Steffy BM, et al. Phosphoregulation of mixed-lineage kinase 1 activity by multiple phosphorylation in the activation loop. Biochemistry. 2004;43:16348–16355.

27. Xu Z, Maroney AC, Dobrzanski P, Kukekov NV, Greene LA. The MLK family mediates c-Jun N-terminal kinase activation in neuronal apoptosis. Mol. Cell. Biol. 2001;21:4713–4724.

28. Fawdar S, Trotter EW, Li Y, Stephenson NL, Hanke F, Marusiak AA, et al. Targeted genetic dependency screen facilitates identification of actionable mutations in FGFR4, MAP3K9, and PAK5 in lung cancer. Proc. Natl. Acad. Sci. U. S. A. 2013;110:12426–12431.

29. Stark MS, Woods SL, Gartside MG, Bonazzi VF, Dutton-Regester K, Aoude LG, et al. Frequent somatic mutations in MAP3K5 and MAP3K9 in metastatic melanoma identified by exome sequencing. Nat. Genet. 2011;44:165–169.

30. Jun L, Xuhong L, Hui L. Circ_SIPA1L1 promotes osteosarcoma progression via miR-379-5p/MAP3K9 axis. Cancer Biother. Radiopharm. 2023;38:604–618.

31. Xia J, Cao T, Ma C, Shi Y, Sun Y, Wang ZP, et al. MiR-7 suppresses tumor progression by directly targeting MAP3K9 in pancreatic cancer. Mol. Ther. Nucleic Acids. 2018;13:121–132.

32. Wu T, Du M, Zhang T, Chen X, Liu Z. Comparisons of global incidence and risk factor profiles of hepatocellular carcinoma and intrahepatic cholangiocarcinoma. Ann. Hepatol. 2025;31:102139.

33. Kozlitina J, Smagris E, Stender S, Nordestgaard BG, Heather H, Tybjærg-hansen A, et al. Exone-wide association study identifies TM6SF2 variant that confers susceptibility to nonalcoholic fatty liver disease. Nat. Genet. 2014;46:352–356.

34. Stender S, Kozlitina J, Nordestgaard BG, Tybjærg-Hansen A, Hobbs HH, Cohen JC. Adiposity amplifies the genetic risk of fatty liver disease conferred by multiple loci. Nat. Genet. 2017;EPub 24 April 2017.

35. Feng G, Targher G, Byrne CD, Yilmaz Y, Wai-Sun Wong V, Adithya Lesmana CR, et al. Global burden of metabolic dysfunction-associated steatotic liver disease, 2010 to 2021. JHEP Rep. 2025;7:101271.

36. Tan DJH, Tamaki N, Kim BK, Wijarnpreecha K, Aboona MB, Faulkner C, et al. Prevalence of low FIB-4 in MASLD-related hepatocellular carcinoma: A multicentre study. Aliment. Pharmacol. Ther. 2025;61:278–285.

37. Tan EY, Danpanichkul P, Yong JN, Yu Z, Tan DJH, Lim WH, et al. Liver cancer in 2021: Global burden of disease study. J. Hepatol. 2024;82:851–860.

38. Sveinbjornsson G, Ulfarsson MO, Thorolfsdottir RB, Jonsson BA, Einarsson E, Gunnlaugsson G, et al. Multiomics study of nonalcoholic fatty liver disease. Nat. Genet. 2022;54:1652–1663.

39. Yang H-C, Kwok P-Y, Li L-H, Liu Y-M, Jong Y-J, Lee K-Y, et al. The Taiwan Precision Medicine Initiative provides a cohort for large-scale studies. Nature. 2025;1–11.

40. Chen H-H, Chen C-H, Hou M-C, Fu Y-C, Li L-H, Chou C-Y, et al. Population-specific polygenic risk scores for people of Han Chinese ancestry. Nature. 2025;1–10.

41. Walters RG, Millwood IY, Lin K, Schmidt Valle D, McDonnell P, Hacker A, et al. Genotyping and population characteristics of the China Kadoorie Biobank. Cell Genom. 2023;3:100361.

42. Sakaue S, Kanai M, Tanigawa Y, Karjalainen J, Kurki M, Koshiba S, et al. A cross-population atlas of genetic associations for 220 human phenotypes. Nat. Genet. 2021;53:1415–1424.

43. Nam K, Kim J, Lee S. Genome-wide study on 72,298 individuals in Korean biobank data for 76 traits. Cell Genom. 2022;2:100189.

44. Machiela MJ, Huang W-Y, Wong W, Berndt SI, Sampson J, De Almeida J, et al. GWAS Explorer: an open-source tool to explore, visualize, and access GWAS summary statistics in the PLCO Atlas. Sci Data. 2023;10:25.

45. Verma A, Huffman JE, Rodriguez A, Conery M, Liu M, Ho Y-L, et al. Diversity and scale: Genetic architecture of 2068 traits in the VA Million Veteran Program. Science. 2024;385:eadj1182.

46. All of Us Research Program Genomics Investigators. Genomic data in the All of Us Research Program. Nature. 2024;627:340–346.

47. All of Us Research Program Investigators, Denny JC, Rutter JL, Goldstein DB, Philippakis A, Smoller JW, et al. The “All of Us” Research Program. N. Engl. J. Med. 2019;381:668–676.

48. Murphy AE, Schilder BM, Skene NG. MungeSumstats: a Bioconductor package for the standardization and quality control of many GWAS summary statistics. Bioinformatics. 2021;37:4593–4596.

49. GitHub - hail-is/hail: Cloud-native genomic dataframes and batch computing [Internet]. GitHub. [cited 2025 Dec 1];Available from: https://github.com/hail-is/hail

50. Mägi R, Horikoshi M, Sofer T, Mahajan A, Kitajima H, Franceschini N, et al. Trans-ethnic meta-regression of genome-wide association studies accounting for ancestry increases power for discovery and improves fine-mapping resolution. Hum. Mol. Genet. 2017;26:3639–3650.

51. Willer CJ, Li Y, Abecasis GR. METAL: fast and efficient meta-analysis of genomewide association scans. Bioinformatics. 2010;26:2190–2191.

52. R Core Team. A language and environment for statistical computing. Vienna, Austria: R Foundation for Statistical Computing. 2019;

53. Watanabe K, Taskesen E, van Bochoven A, Posthuma D. Functional mapping and annotation of genetic associations with FUMA. Nat. Commun. 2017;8:1826.

54. Watanabe K, Umićević Mirkov M, de Leeuw CA, van den Heuvel MP, Posthuma D. Genetic mapping of cell type specificity for complex traits. Nat. Commun. 2019;10:3222.

55. 55. GitHub - MRCIEU/gwasglue: Linking GWAS data to analytical tools in R [Internet]. GitHub. [cited 2025 Dec 1];Available from: https://github.com/MRCIEU/gwasglue

56. Kim JJ, Vitale D, Otani DV, Lian MM, Heilbron K, 23andMe Research Team, et al. Multi-ancestry genome-wide association meta-analysis of Parkinson’s disease. Nat. Genet. 2024;56:27–36.

57. McLaren W, Gil L, Hunt SE, Riat HS, Ritchie GRS, Thormann A, et al. The Ensembl Variant Effect Predictor. Genome Biol. 2016;17:122.

58. Gusev A, Ko A, Shi H, Bhatia G, Chung W, Penninx BWJH, et al. Integrative approaches for large-scale transcriptome-wide association studies. Nat. Genet. 2016;48:245–252.

59. Grishin D, Gusev A. Allelic imbalance of chromatin accessibility in cancer identifies candidate causal risk variants and their mechanisms. Nat. Genet. 2022;54:837–849.

60. Liu Y, Basty N, Whitcher B, Bell JD, Sorokin EP, van Bruggen N, et al. Genetic architecture of 11 organ traits derived from abdominal MRI using deep learning. Elife [Internet]. 2021;10. Available from: 10.7554/eLife.65554

61. Pulit SL, Stoneman C, Morris AP, Wood AR, Glastonbury CA, Tyrrell J, et al. Meta-Analysis of genome-wide association studies for body fat distribution in 694 649 individuals of European ancestry. Hum. Mol. Genet. 2019;28:166–174.

62. Huerta-Chagoya A, Schroeder P, Mandla R, Li J, Morris L, Vora M, et al. Rare variant analyses in 51,256 type 2 diabetes cases and 370,487 controls reveal the pathogenicity spectrum of monogenic diabetes genes. Nat. Genet. 2024;56:2370–2379.

63. Rashkin SR, Graff RE, Kachuri L, Thai KK, Alexeeff SE, Blatchins MA, et al. Pan-cancer study detects genetic risk variants and shared genetic basis in two large cohorts. Nat Commun [Internet]. 2020;11. Available from: 10.1038/s41467-020-18246-6

64. Fernandez-Rozadilla C, Timofeeva M, Chen Z, Law P, Thomas M, Schmit S, et al. Deciphering colorectal cancer genetics through multi-omic analysis of 100,204 cases and 154,587 controls of European and east Asian ancestries. Nat. Genet. 2023;55:89–99.

